# Airborne particulate matter enhances with monosodium urate crystals the secretion of IL-1β by human immune cells

**DOI:** 10.64898/2026.02.26.26347218

**Authors:** Atefeh Razazan, Marilyn E Merriman, Nandi Burden, Richard J Reynolds, Leo AB Joosten, Salik Hussain, Tony R Merriman

## Abstract

Gout is driven by an interleukin-1*β*-mediated intense innate immune reaction to monosodium urate (MSU) crystals (MSUc). In cell culture models of inflammatory gout there is a synergistic effect of phagocytosis of MSUc and TLR2 and TLR4 activation by agonists such as free fatty acid and lipopolysaccharide (LPS) in NLRP3-inflammasome activation and IL-1*β* secretion. A substantial number of gout patients do not report a dietary trigger, and observational studies associate airborne particulate matter with incident gout and flares. Airborne particulate matter contains LPS and airborne-derived particulate matter stimulates IL-1*β* secretion in cell culture. We hypothesized that air-borne particulate matter could co-stimulate, with MSUc, IL-1*β* secretion and inflammation. We tested the hypothesis using MSUc with extracted airborne PM_4_ in human cells (the THP-1 monocyte cell line, primary human monocytes and PBMCs) or carbon black particles with ozone (CB+O_3_) in a murine foot-pad injection model of gout. There was strong NLRP3-inflammasome-dependent co-stimulation of IL-1*β* secretion in THP-1 cells with PM_4_+MSUc and a moderate additive effect in primary human PBMCs. However, there was no added effect on IL-1*β* secretion of PM_4_ in isolated primary human monocytes. Inhalation of CB+O_3_ persistently exacerbated MSUc-induced murine paw inflammation, with an increase of alveolar/lavage macrophages that contained CB+O_3_ particles and increased lavage expression of IL-1*β*. In conclusion, airborne-derived PM_4_ particulate matter enhanced MSUc-induced IL-1*β* secretion in THP-1 cells and PBMCs. Combined with exacerbation of MSUc-induced inflammation by fine particulate matter in *in vivo* experiments, these data provide evidence that exposure to fine particulate matter may play a role in the etiology of gout.

## Introduction

Gout is caused by an intense innate immune response to monosodium urate (MSU) crystals (MSUc) deposited in the synovial cavity due to hyperuricemia^1^. Macrophages (differentiated from circulating monocytes) are primary effector cells. The central molecular mechanism, secretion of bioactive IL-1*β* is governed by activation of the NLRP3 inflammasome in response to each of signaling through the toll-like 2 and 4 receptors and phagocytosis of MSUc^2^. In cell culture models, either of MSUc or toll-like receptor (TLR) agonist (e.g. diet-derived free fatty acid for TLR2^3^ or lipopolysaccharide (LPS, an endotoxin) for TLR4) elicit a relatively modest IL-1*β* response from monocytes with a very synergistic response observed in the presence of both MSU crystal and agonist^3, 4^.

The presence of MSUc in asymptomatic hyperuricemia is common (for example 34%^5^), indicating the necessity of additional factors to progress to gouty inflammation. Genetic variation in genes involved in the regulation of NPRP3-inflammasome formation and activity have been identified^6^, and the only proven environmental factors are certain foods that trigger a gout flare (for example red meat, seafood or alcohol^7, 8^). However, a majority of people with gout in the UK (63%) and 42% of European people in New Zealand report no known food trigger^9, 10^ .

Air pollution is a serious public health issue. Exposure increases the risk of cardiometabolic disease, contributing to the burden of disease attributable to environmental exposures^11, 12^. There is some, but not compelling, evidence from observational epidemiology for a role of airborne particulates in gout: a prospective study in the UK associated a 3% risk of incident gout with each interquartile increase in exposure to particulate matter of 2.5 microns (PM_2.5_) ^13^; a second study in the UK Biobank studied prevalent gout, associating a 5% increase in risk with each interquartile increase in exposure to PM_2.5_^14^; exposure to PM_2.5_ in Taiwan associated with a 44% increased risk of prevalent gout comparing the top quartile of exposure to the bottom quartile^15^; and a Korean time-series study associated short-term exposure to PM_10_ with increased risk of emergency department visits attributable to a gout flare^16^. We note, however, in the Liu et al. ^13^ incident gout study only 398 people with prevalent gout were identified at baseline, a gross underestimate of the true number of prevalent cases^17^.

There is considerable evidence from cell culture models that airborne-derived particulates can stimulate the NLRP3 inflammasome, for example: PM_10_ induced an ∼25-fold increase in IL-1*β* secretion in human peripheral blood mononuclear cells (PBMCs) ^18^; PM_10_ collected from various sites in Quito, Ecuador stimulated IL-1*β* secretion from mouse bone marrow-derived macrophages when cells were primed with LPS^19^; PM_10_ from the National Institute of Standards and Technology (NIST) stimulated IL-1*β* secretion in human monocytes^20^; and PM_10_ collected from Milano, Italy stimulated IL-1*β* secretion in phorbol myristate acetate-differentiated THP-1 cells^21^. Moreover, exposing mice to fine airborne particulate matter increases IL-1*β* levels in bronchoalveolar lavage^22-25^.

Airborne fine particulate matter contains LPS. Galuszka et al. ^26^ quantitated LPS in NIST PM_6_ to be 1.75 endotoxin units (or ∼0.25ng) per *μ*g of particulate matter. Removal of LPS from the particulate matter reduced the inflammatory response of primary human monocytes^26^. This is consistent with the hypothesis that fine airborne particulate matter is a vehicle for delivery of the TLR4-agonist LPS to monocytes. In the context of gout, it is possible that fine airborne particulates co-activate with MSU crystals the NLRP3 inflammasome. Here we tested this hypothesis using THP-1 cells, primary monocytes, peripheral blood mononuclear cells and a murine model of gout.

## Reagents and Methods

### Reagents

Environmental airborne particulate matter was purchased from NIST (Standard Reference Material 2786), with mean diameter > 4μm (PM_4_). These particles were collected in 2005 from the Nuclear Physics Institute in Prague, Czech Republic. The PM_4_ was resuspended in Roswell Park Memorial Institute (RPMI) medium and used at a final concentration of 100 *μ*g/ml, with sonication before use in a Fisher FS30 40KHz 130W water bath for 5-min. LPS (*E. coli* serotype 055:B5) was purchased from Sigma and was used at a final concentration of 1 ng/ml. MSUc was purchased from Invivogen and used at a final concentration of 300 *μ*g/ml. The carbon black (CB) was a gift from Evonik, Frankfurt, Germany. Sterile CB+O_3_ was generated using an inhalation exposure system by aerosolizing CB in the presence of O_3_ as described in detail in ref^24^ and was used at a concentration of 100 *μ*g/ml in the THP-1 stimulation. The average size of the CB_OX_ was 80 nm. NLRP3-inflammasome inhibitor MCC950 was purchased from Invivogen and used at 2 nmol/l.

### Cell culture

THP-1 cells were obtained from the American Type Culture Collection (Manassas, VA) and cultured in RPMI 1640 media (Gibco, Thermo Fisher, Waltham, MA) supplemented with 10% fetal bovine serum, with 5×10^-5^ cells added to each stimulation. Peripheral blood mononuclear cells (PBMCs) from healthy donors were prepared from Cell Preparation Tubes (CPT) containing Ficoll with red blood cells lysed using cold ammonium-chloride-potassium buffer. Monocytes, prepared from PBMCs prepared from healthy donors using the Stem Cell Technologies (Cambridge, MA) EasySep™ Human Monocyte Isolation kit, were cultured in RPMI 1640 media supplemented with 10% human pooled serum with 1×10^5^ cells added to each stimulation. THP-1 and PBMC stimulations were carried out for 24-hr and for human primary monocytes stimulations were done for 16-hr. THP-1 cells were primed for one-hour with LPS before adding CB_OX_ particles. Extracellular IL-1*β* was quantified by enzyme-linked immunosorbent assay (R&D Systems, Minneapolis, MN).

### Murine model

Eight-week old C57BL/6J female mice were obtained from Jackson Laboratory (Bar Harbor, ME) and maintained for one-week in the West Virginia University animal care facility before exposure. The animals were maintained with a 12-hour light / dark cycle and were provided chow and water *ad libitum*. MSUc (5mg/kg intradermal; Invivogen) were injected in the left hind paw, with right hind paw phosphate-buffered saline (PBS) injection as a control. Hind paw thickness was measured to establish the severity of tarsal/ankle oedema. A thickness index/swelling ratio was calculated (Δ circumference at various time points-circumference at time 0)/circumference at time 0) ^27^. Whole-body inhalation co-exposure to the CB+O_3_ (2.5 mg/m^3^CB+ 1ppm O_3_) was performed on days 9 and 10 post MSUc injection as described in ref^23^. Euthanasia was carried out by intraperitoneal injection of Fatal Plus (250 mg/kg) 24-hour post exposure to particles. The West Virginia University Institutional Animal Care and Use Committee, an AALAC-accredited program, approved the animal procedures.

### Quantification of lung inflammation and injury, particle uptake by macrophages, IL-1β gene and protein expression

Bronchoalveolar lavage was performed immediately after euthanasia on the right lung lobes of mice, and the unlavaged left lung lobe was snap frozen in liquid nitrogen and stored at -80°C until further use^28^. Ice-cold PBS (0.5 mL) was instilled three times through the trachea, and the collected fluid was used for total cell counts in the lavage. Bronchoalveolar lavage fluid was spun at 400g for 5 minutes, and the cell pellet was resuspended in PBS. Resuspended cells were spun on a slide using Cytspin4 (ThermoFischer Scientific), and slides were stained with Hema 3 (Fisher Scientific). A differential cell count was performed to quantify macrophage and neutrophil populations. These slides were also used to calculate the number of particle-positive macrophages which was assessed by quantifying the percentage of CB (black aggregates)-positive lavage macrophages as previously described^25^. The Hema 3-stained slides were imaged using Olympus AX70 Microscope (Japan) at 40X magnification. IL-1□ gene expression in lung tissue and protein release in the bronchoalveolar lavage fluid was performed by real-time PCR and Duoset ELISA as described previously^25^.

### Statistical analysis

In the cell culture experiments we tested the hypothesis that PM_4_ and MSUc are synergistic in stimulating IL-1*β* production. Factorial designs were carried out combining pairs of PM_4_, MSUc and LPS in replicate samples. The statistical models were performed separately for the three cell types, THP-1, PBMCs and monocytes. A linear mixed model with treatments as fixed main effects and their interactions and with a biological replicate-specific intercept as a random effect was fit to the IL-1*β* concentration data. The null hypothesis of additivity in pairwise combinations of the three stimuli was rejected when the respective two-way interaction effect had P < 0.05. The IL-1*β* data were positively skewed, therefore the data were natural log transformed (THP-1 cells and primary monocytes) or square root transformed (PBMCs) to minimize outlier effects (very large estimated concentrations of IL-1*β*) and to meet normality assumptions of the linear models. P-values of the interaction effects were obtained by loglikelihood ratio tests of the full model compared to a reduced model, absent the interaction effect in question. The resulting Chi square statistic had degrees of freedom equal to one in all cases. IL-1*β* estimated marginal means and confidence limits for stimulus treatment combinations were obtained using the emmeans function in the R package, emmeans ^29^. The linear mixed models were fit using the function lmer in the R package lme ^30^. Statistical programming was performed in R version 4.5.2 ^31^ .

Animal study data are presented as mean ± standard error of mean and were generated using 5-8 mice per group. Data were analyzed by one-way analysis of variance (ANOVA) followed by Tukey’s multiple comparisons post hoc test. Two group comparisons were made using an unpaired Student’s t-test.

## Results

### Co-stimulation of IL-1β secretion by MSU crystals and PM_4_ in THP-1 cells, human primary monocytes and peripheral blood mononuclear cells (PBMCs)

Data presented in **Figure 1** demonstrate that PM_4_ non-additively co-induces with MSU crystals (MSUc) IL-1*β* secretion by THP-1 cells (*P*_MSUc*PM4_ = 0.026) – an average of 4199 pg/ml of IL-1*β* was secreted in the co-stimulations compared to 306 pg/ml for MSUc alone and 468 pg/ml for PM_4_ alone. In the same experiments LPS also non-additively co-induced with MSUc IL-1*β* secretion (*P*_MSUc*LPS_ = 0.011) – an average of 1322 pg/ml of IL-1*β* was secreted in the co-stimulations compared to 306 pg/ml for MSUc alone and 225 pg/ml for PM_4_ alone. The co-stimulation with MSUc of IL-1*β* secretion by PM_4_ in the THP-1 cells was dependent on NLRP3 inflammasome activity, with IL-1*β* concentration greatly reduced with the addition of NLRP3 inflammasome inhibitor MCC950 - in a single experiment with two technical replicates IL-1*β* concentration reduced from 9734 pg/ml to 485 pg/ml (when LPS and MSUc were used together the IL-1*β* concentration reduced from 1747 pg/ml to 154 pg/ml in the presence of MCC950). However, in human primary monocytes there was no evidence of non-additive interaction between MSUc and PM_4_ in co-inducing IL-1*β* secretion (*P*_MSUc*PM4_ = 0.026; an average of 3723 pg/ml of IL-1*β* was secreted in the co-stimulation compared to 0 pg/ml for MSUc alone and 3603 pg/ml for PM_4_ alone) (**Figure 2**). In contrast, there was non-additive co-stimulation with MSUc and LPS in the primary monocytes (*P*_MSUc*LPS_ = 0.0023). In PBMCs, while there was an increase in IL-1*β* secretion in cells co-stimulated with MSUc and PM_4_ compared to PM_4_ alone (two-tailed paired t-test *P* = 0.046), there was no statistical evidence for non-additive co-stimulation (*P*_MSUc*PM4_ = 0.16) whereas there was for MSUc and LPS (*P*_MSUc*LPS_ = 3.3×10^-8^) (**Figure 3**).

**Figure 1.**
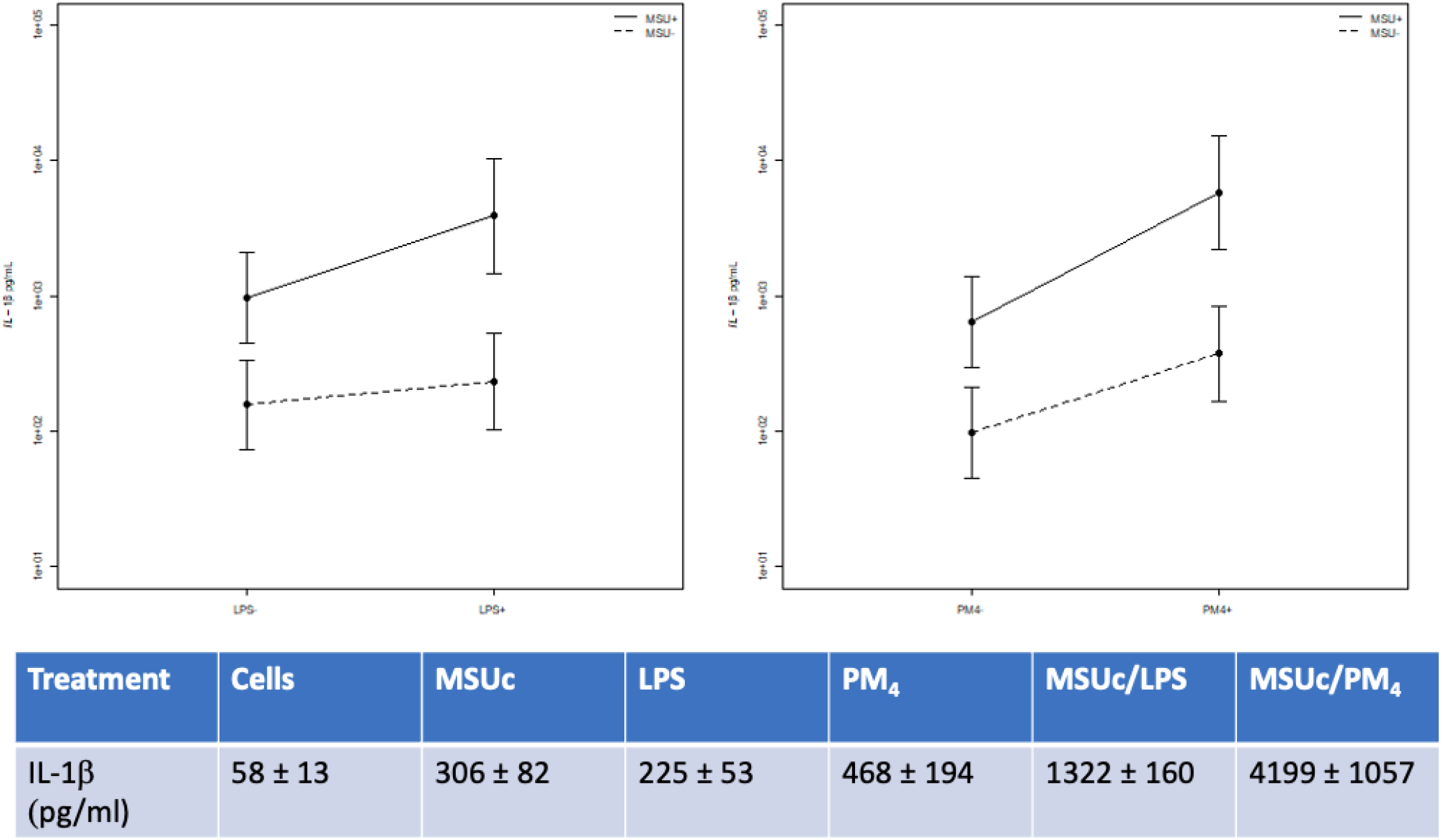
Stimulation of THP-1 cells. The plots show, on the left, stimulations with MSUc ± LPS and, on the right, with MSUc ± PM_4_. The table shows the average IL-1*β* concentrations ± standard error of the mean for each treatment. The data are derived from stimulations done on five separate days with two technical replicates per treatment.

**Figure 2.**
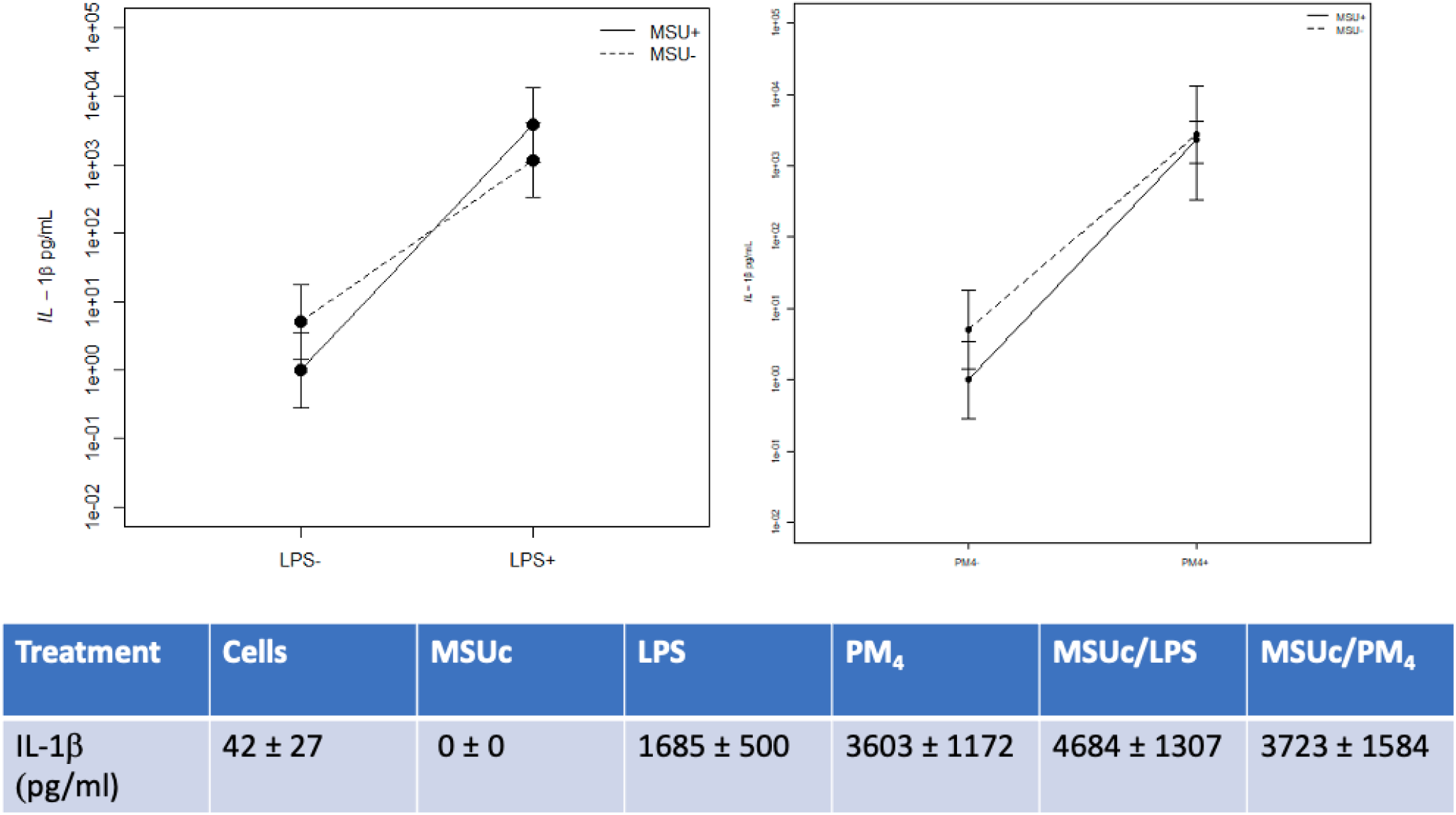
Stimulation of primary monocytes. The plots show, on the left, stimulations with MSUc ± LPS and, on the right, with MSUc ± PM_4_. The table shows the average IL-1*β* concentrations ± standard error of the mean for each treatment. The data are derived from stimulations done on three separate days from three separate donors with two technical replicates per treatment.

**Figure 3.**
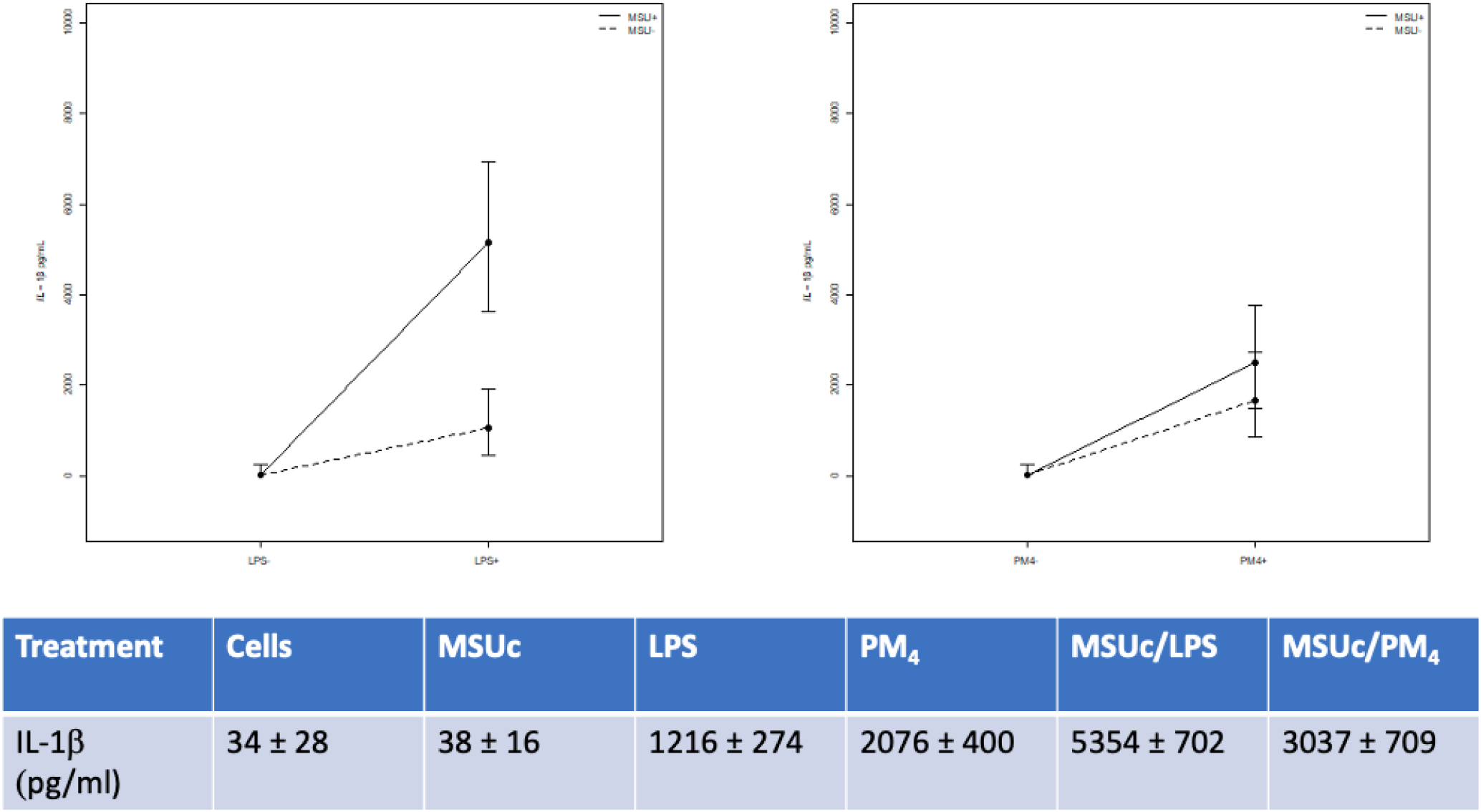
Stimulation of PBMCs. The plots show, on the left, stimulations with MSUc ± LPS and, on the right, with MSUc ± PM_4_. The table shows the average IL-1*β* concentrations ± standard error of the mean for each treatment. The data are derived from stimulations done on seven separate days from seven separate donors with two technical replicates per treatment.

### Exacerbation of footpad swelling in mice with MSU crystal injection and CB_OX_ exposure associates with a pulmonary inflammatory phenotype

In the *in vivo* experiments presented here, owing to cost, animals were exposed to CB+O_3_ instead of PM_4_. To verify that these particulates behaved similarly to PM_4_ in the THP-1 cells we stimulated THP-1 cells with CB+O_3_ primed with LPS. In a single experiment with two technical replicates LPS + CB+O_3_ induced secretion of 1107 pg/ml of IL-1*β* and, with the addition of MSUc to the LPS + CB+O_3_, 3783 pg/ml of IL-1*β* was secreted. When MSUc was injected into the footpad of mice, inhalation of CB+O_3_ persistently exacerbated swelling at day 10 (paw swelling index was 0.18 with exposure and 0.11 without) (**Figure 4**). Exposure to the CB+O_3_ particles associated, in the resolving phase of the MSUc-induced paw inflammation, with induction of a pulmonary inflammatory phenotype (increased lavage neutrophils and macrophages), which was exacerbated by paw injection of MSUc (**Figure 5A,B**). There was a significant increase in the number of alveolar/lavage macrophages that contained CB+O_3_ particles (**Figure 5C**). Increased lavage IL-1*β* mRNA and protein expression levels were observed with MSUc paw injection with and without CB+O_3_ exposure (**Figure 5D,E**).

**Figure 4.**
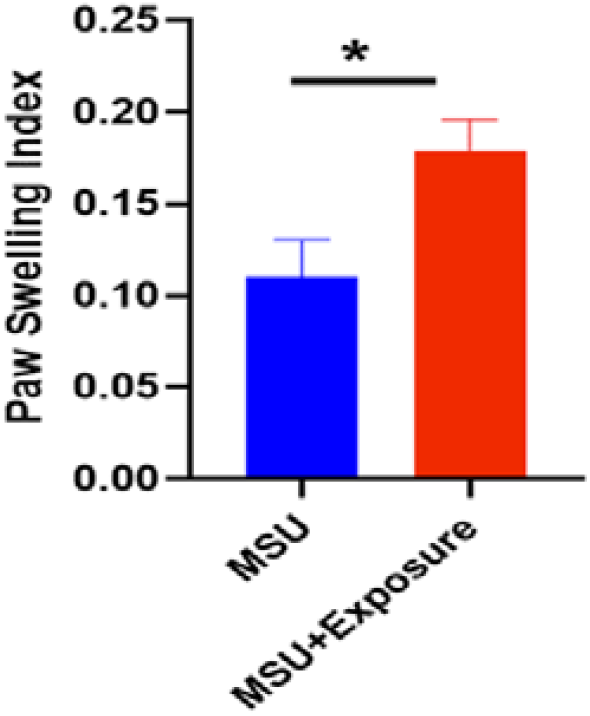
Persistence of paw swelling by inhalation exposure of CB+O_3_ day 9,10 post MSUc injection. N= 4-5 per group. \**P*<0.05.

**Figure 5.**
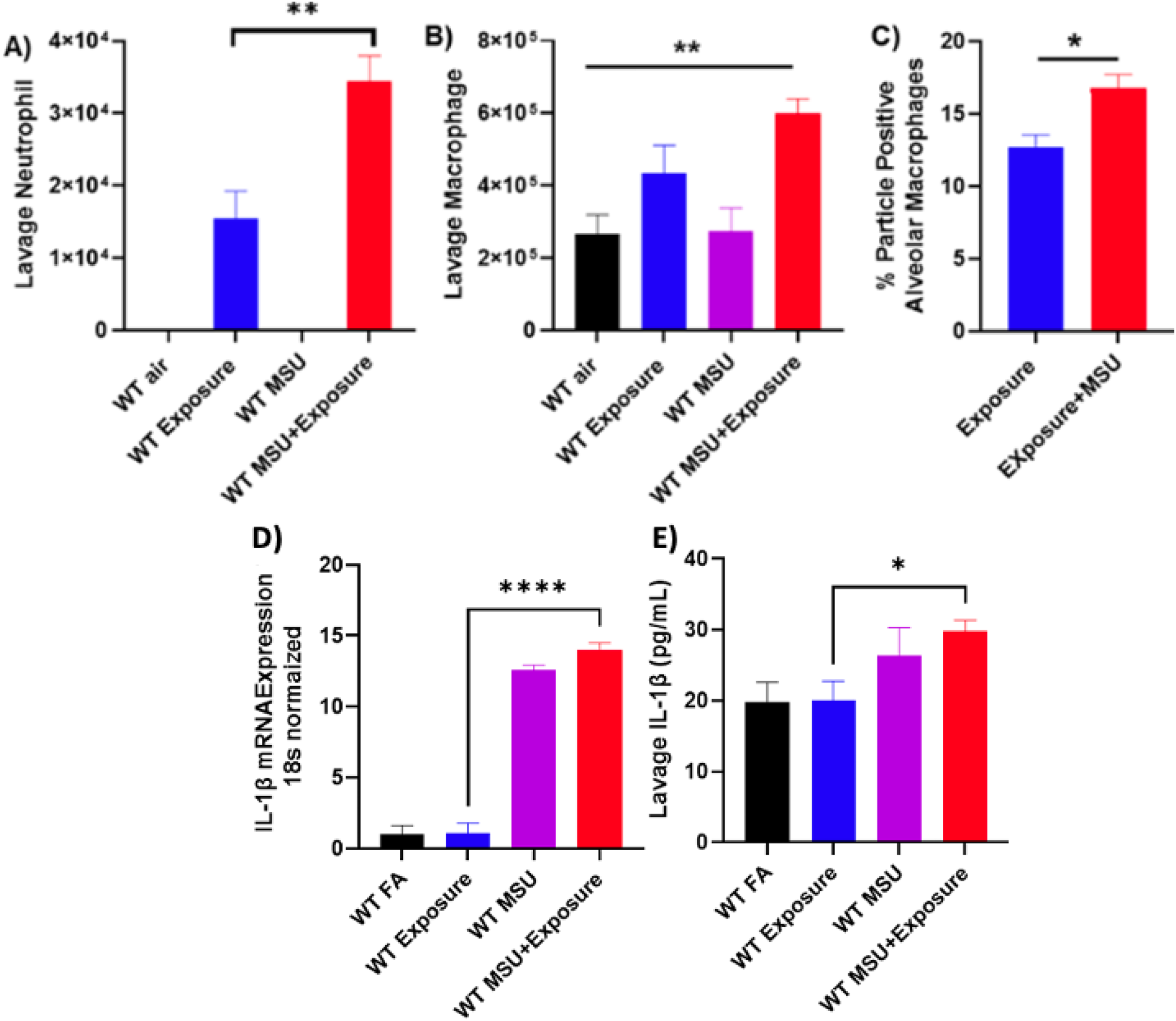
Innate immune cell phenotype and IL-1*β* expression after exposure of animals to CB+O_3_. Absolute counts for A) lavage neutrophils B) macrophages and C) particle-containing macrophages in lavage, n=4-6 per experiment. Evidence of NLRP3-inflammasome activation in the lungs after MSU injection and air pollution exposure day 8 post MSUc injection (D) pro-IL-1□ mRNA expression and E) IL-1□ concentration in lung lavage fluid, n=4-6 per experiment. \**P*<0.05, ** *P*<0.005, **** *P*<0.00005.

## Discussion

Triggering of a gout flare requires co-stimuli that act on innate immune cells. The current orthodoxy is that these stimuli are phagocytosis of MSUc and activation of TLR2 and / or TLR4. Bacterial endotoxin (LPS) activates TLR4 and airborne fine particulate matter contains LPS^26^. In cell culture, airborne particulate matter can activate IL-1*β* secretion from PBMCs, monocytes and macrophages^18-21^, there is increased IL-1*β* in the bronchoalveolar lavage of mice exposed to fine particulate matter^22-25^, and epidemiological studies associate exposure to airborne pollution with incident and prevalent gout and the occurrence of flares^13-16^. We hypothesized that airborne fine particulate matter could be an environmental exposure that co-induces with MSUc IL-1*β* secretion in innate immune cells. We tested this hypothesis in cell culture experiments and a murine model of acute gouty arthritis and found that PM_4_ could co-stimulate with MSU crystal IL-1*β* secretion in an NLRP3-inflammasome-dependent manner in the THP-1 monocyte cell-line in a similar manner to LPS. However, in primary human cells co-stimulation was either absent (monocytes) or less effective than with LPS (PMBCs). Furthermore, exposure of mice to airborne CB+O_3_ particles exacerbated footpad inflammation caused by MSUc injection. The murine data demonstrate that fine particulate matter can exacerbate MSUc-induced inflammation *in vivo*. Collectively, our findings support the hypothesis that airborne fine particulate matter has a causal role in gout, possibly by acting as a bacterial endotoxin delivery vehicle to TLR4. Our experimental data support the increasing epidemiological evidence for a role of airborne fine particulates in gout.

In primary human circulating white blood cells the results were inconsistent. In monocytes there was no additional effect of MSUc on IL-1*β* secretion over the effect of PM_4_ alone. This can be partly attributed to the phenomenon of freshly isolated human primary monocytes constitutively expressing active caspase-1^32^, meaning that less NLRP3-inflammasome activation is required. However, in PBMCs there was an ∼1.5-fold increase in IL-1*β* secretion when MSUc was included with PM_4_. The differences in response between the THP-1 cell-line and human primary cells was notable. The THP-1 cell line is an immortalized monocyte-like cell line derived from a child with acute monocytic leukemia. In THP-1 cells there was an ∼10-fold increase in IL-1*β* secretion when MSU crystals were included with PM_4_, compared to no increase in primary monocytes and an ∼1.5-fold increase in PBMCs. Consistent with previous data^33^ THP-1 cells in our experiments had a lesser IL-1*β* response to LPS than primary monocytes which is correlated to lower CD14 expression in THP-1 cells^34^. There was a similar pattern in the IL-1*β* response to PM_4_. Other differences between THP-1 cells and primary monocytes include substantial differences in chromatin structure and transcriptional activity that leads to a lesser immune cell phenotype^35^. Collectively the experiments presented in **Figure 1** do provide some support for a role of airborne particulates in stimulating a NLRP3-inflammasome response in the presence of MSUc, however the data do have to be interpreted understanding that THP-1 cells are an imperfect clinical model, that no role was demonstrated in primary monocytes and a modest role in PBMCs.

A role for exposure to inhaled particulates in amplifying innate response to MSUc was supported in the *in vivo* model. It was intriguing that, during exposure to CB+O_3_ particles, MSUc injection in the food pad led to increased alveolar macrophage and neutrophil count, and an increased number of alveolar macrophages having ingested CB+O_3_ particles. Furthermore, increased IL-1*β* mRNA and protein expression was observed in lavage upon foot pad MSU crystal injection, irrespective of CB+O_3_ particulate exposure. These findings suggest systemic immune changes that are induced after MSUc injection in the foot pad that lead to alterations in pulmonary inflammatory response. They also suggest communication between the joints and the lung in the inflammatory response upon MSU crystal and particulate exposure. There is evidence for differential response of resident peritoneal immune cells, at least, to various footpad stimuli^36^.

There is an incontrovertible close temporal relationship between ingestion of certain foods as triggers of gout flares^7, 8^. Given that this relationship has long been established, it follows that research into environmental exposures and gout has heavily focused in the previous 2-3 decades on the role of diet, in particular the association of individual dietary components with urate concentration and risk of gout. These associations, and overall dietary patterns have a small effect size on urate level and risk of gout and are at least partly mediated by BMI^37, 38^. In contrast there is relatively little epidemiological literature on other environmental exposures, including particulates. While there is some epidemiological evidence for airborne pollution as a cause of gout, we are unaware of any studies examining other types of particulate matter. Candidate types of particulate matter include titanium-based particulates, micro- and nano-plastics, and orthopedic wear debris. All have been demonstrated to induce IL-1*β* secretion in human primary monocytes and THP-1 cells^39-41^. Titanium oxide particles, for example, are present in synovial fluid and their prevalence in synovial fluid in Dutch rheumatology patients dropped after the European Union banned their use in food products in early 2022^42^. Whether or not they, and other aforementioned particulates, co-stimulate IL-1*β* secretion with MSUc, analogous to what we observed with PM_4_, is unknown.

These studies were conducted at pollutant exposure levels anticipated to be deposited within a short time frame. Normally, the exposure to air pollution is lifelong and overall deposition depends upon the particulate matter characteristics (size, shape, concentration), physiological considerations (uptake and clearance by the alveolar macrophages) and co-existing conditions (chronic respiratory diseases etc). We previously published the deposition of CB+O_3_ aerosol at various concentrations in the mouse lung^23-25, 43^. Pulmonary deposition after a single 3-hour exposure at 1mg/m^3^ CB+O_3_is equivalent to an anticipated deposited dose from a 3.5-day exposure at the current national ambient air quality standard (NAAQS) for PM_2.5_. These levels are relevant for environmental exposures as global PM_2.5_ average levels routinely exceed NAAQS for approximately 99% of the urban population^44^. Anatomical differences and the sedentary nature of laboratory rodents necessitate exposure at 4-5 times than for human to demonstrate similar biological outcomes^45, 46^. O_3_ exposure at 1 ppm is 2.5-3.1 times the current NAAQS multiplied by 4-5 and is relevant to human clinical trial O_3_ exposure levels of 200-400 ppb (2.5-5 times) ^47-49^.

In summary, these findings highlight the role of air pollution as a co-stimulant of MSUc-induced NLRP3 inflammasome activation in experimental gouty arthritis. Further research could include chronic and even lower concentration air pollution exposures, evaluation of other particulates in the experimental models used herein, and examination of molecular mediating mechanisms.

## Data Availability

All data produced in the present work are contained in the manuscript.

## Acknowledgements

The authors would like to thank the people who donated blood samples for this study as part of the University of Alabama at Birmingham Healthy Donor Cohort. The National Institute of Health is acknowledged for funding (R01ES031253).

